# Playing position and long-term mortality among elite male football players, 1930–1990

**DOI:** 10.64898/2026.02.16.26346414

**Authors:** Dirk Witteveen, David K. Humphreys

**Author notes:** Correspondence to: Dirk Witteveen.

## Abstract

**Background:** Concern about long-term health effects of repetitive head impacts in football has increased, but it remains unclear whether position-specific exposure patterns were associated with differential long-term all-cause mortality among elite players across the 20th century.

**Methods:** We conducted two retrospective cohort studies of elite male professional football players. The World Cup cohort included all players on the team rosters from FIFA World Cup tournaments (1930–1990), and the UEFA European Cup cohort included all players who appeared in annual quarterfinal, semifinal, or final matches (1956–1991). Vital status was ascertained through archival linkage. Playing position was harmonized into six categories. Age was the time scale. Cox proportional hazards models were stratified by birth cohort and adjusted for origin region; interaction models were used to estimate region-specific marginal hazard ratios.

**Findings:** The World Cup cohort included 4,223 players (2,330 deaths), and the European Cup cohort included 2,710 players (1,126 deaths). In the World Cup cohort, goalkeepers had lower mortality than midfielders (hazard ratio [HR] 0.73, 95% CI 0.63–0.84), whereas center-forwards had higher mortality (HR 1.27, 95% CI 1.08–1.50); mortality among center-backs was elevated but not statistically significant (HR 1.13, 95% CI 0.98–1.31). In the European Cup cohort, center-backs (HR 1.28, 95% CI 1.07–1.55) and other defenders (HR 1.20, 95% CI 1.02–1.42) had higher mortality than midfielders. Region-stratified marginal estimates indicated that elevated risks for central playing roles were greatest in Northwestern Europe and Central/Eastern Europe.

**Interpretation:** Among footballers active during the 20th century, long-term all-cause mortality differed by playing position and varied by region, with higher risks concentrated in central attacking and defensive roles. These patterns were most pronounced in regions where aerial contests historically predominated, suggesting that long-term health risks associated with professional football participation vary by role-specific exposure profiles.

## Introduction

Growing concern surrounds the long-term neurological risks associated with repetitive head impacts in football, particularly those resulting from heading the ball^1^. Football is unique among major team sports in that it involves routine, intentional use of the unprotected head to strike or redirect an airborne ball^2^. As a result, professional players may perform many of thousands of headers in training and games over the course of their careers^3^, which has been associated with cognitive impairment in retrospective cohorts^4^. The heightened awareness of neurocognitive sequelae in contact-sport athletes has prompted precautionary responses from governing bodies, including restrictions on heading during youth training.

At the population level, epidemiological studies link professional football to elevated risks of neurodegenerative disease and mortality. For example, a cohort study of former Scottish professionals found more than threefold higher neurodegenerative mortality compared to the general population, alongside lower all-cause mortality through midlife driven by fewer cardiovascular and cancer deaths up to age 70^5,6^. Moreover, Swedish top-division players had higher dementia incidence than matched controls, while overall survival remained modestly improved^7^. Excess dementia risk was confined to outfield players, whereas goalkeepers exhibited no increased risk, implicating exposures specific to field play. Similarly, a study of English players found that defenders, but not goalkeepers, experienced particularly elevated neurodegenerative mortality^8^, suggesting that long-term health risks in professional football may vary meaningfully within the sport.

One plausible explanation for this heterogeneity is the systematic variation in exposure to head impacts across playing positions. Football players occupy distinct roles that entail position-specific training, tactical responsibilities, and physical demands. These roles shape the frequency of strategic heading – including to pass the ball, score goals, or clear the ball defensively^2,3^ – typically involving repetitive ball-to-head impacts. Strategic heading frequently occurs in the context of aerial contests, in which players challenge opponents for airborne balls under pressure and at speed, adding exposure to accidental head-to-head and elbow-to-head collisions. Such collisions occur disproportionately in defensive play^2,9–11^. In addition, not all headers are biomechanically equivalent: pressured clearances of long goal kicks or crosses involve higher ball velocities (70-85 km/h) and impart substantially greater forces than uncontested headers^12,13^. Playing position therefore serves as a proxy for both the frequency and intensity of exposure to *head impacts*, with potential implications for long-term disease and mortality risk.

Figure 1 illustrates a stylized tactical football formation distinguishing six analytic playing positions: a goalkeeper, center-backs, other defenders, midfielders, center-forwards, and other forwards. Center-backs and center-forwards are consistently more involved in headers and aerial contests than other positions and thus experience greater cumulative exposure to head impacts, motivating the hypothesis that these positions may be at elevated risk of premature mortality. Although tactical systems and position labels evolved over the 20th century, these central defensive and attacking roles remained functionally stable with respect to headers and aerial play. We therefore classify players into harmonized analytic positions designed to capture historically persistent differences in exposure to head impacts.

**Figure 1.**
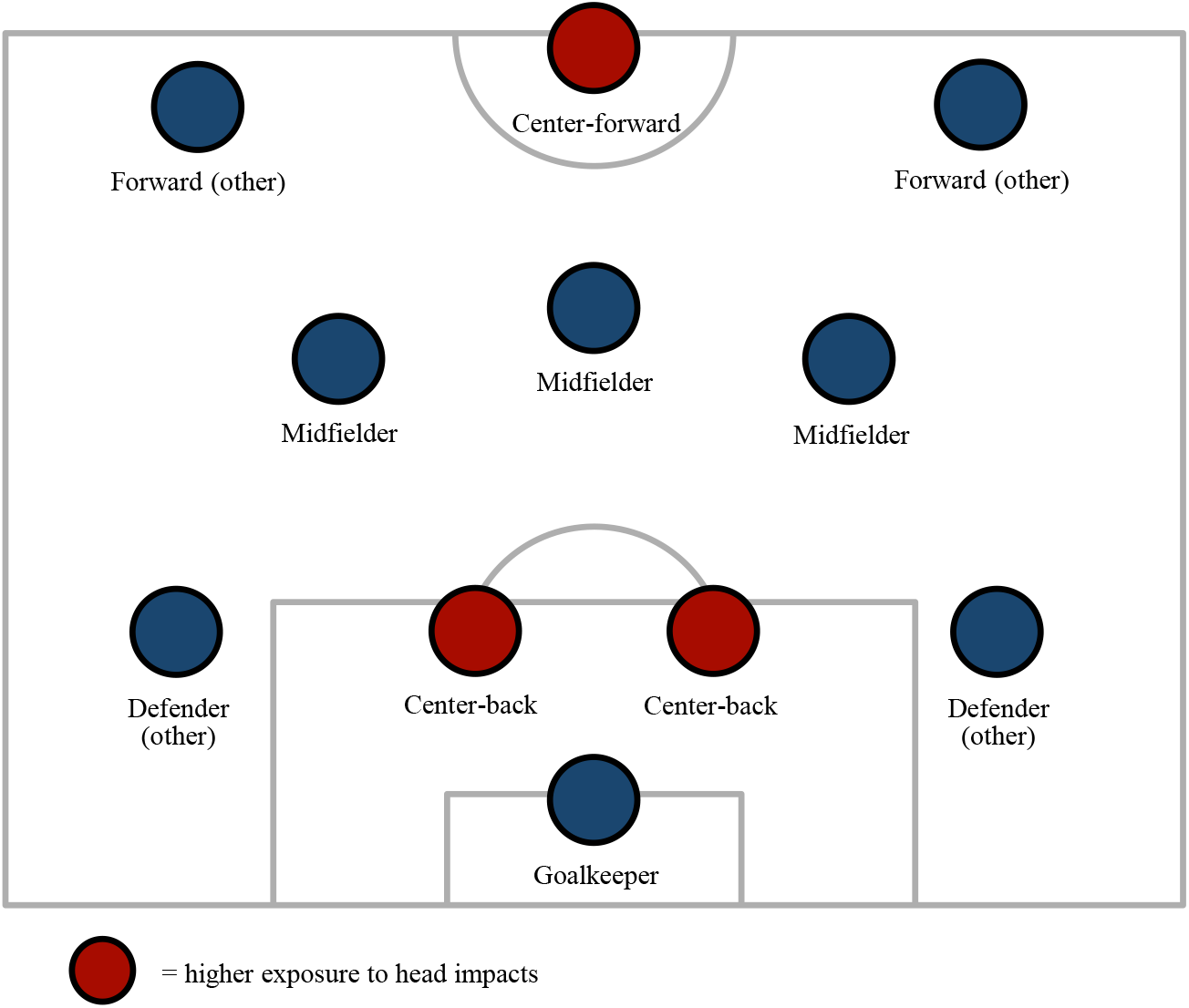
Stylized tactical football formation distinguishing six analytic playing positions. The figure shows the six analytic playing positions used in the study: goalkeeper, center-back, defender (other), midfielder, center-forward, and forward (other). Center-backs and center-forwards are shown in central defensive and attacking roles that are typically involved in aerial contests. Detailed position coding and historical harmonization are described in appendix p 2.

Importantly, positional exposure has historically interacted with regional playing styles. During much of the 20th century, teams in Northwestern and Eastern Europe often favored direct styles characterized by frequent long balls and aerial contests, whereas Southern European and South American traditions emphasized shorter passing and ground-based play^14,15^. Climatic conditions and poorly maintained playing surfaces in Northern parts of Europe may have limited the effectiveness of ground-based passing for much of the 20th century, encouraging more direct styles of play that involve more long balls and aerial challenges. As a result, players occupying central defensive and attacking roles in certain regions may have accumulated greater lifetime exposure to head impacts than their counterparts elsewhere.

Despite growing evidence linking professional football participation to neurodegenerative disease, critical gaps remain. Existing studies have largely focused on players active in the late 20th century or later and have rarely examined long-term mortality differences across specific playing positions in earlier eras. It remains unknown whether position-specific exposure patterns translated into differential all-cause mortality among football players competing during periods when heading and aerial play were prevalent. The primary objective of this study was to examine whether long-term mortality risk differed by playing position among elite male football players participating in major international and club competitions during the majority of the 20th century.

## Methods

### Study design and data sources

We conducted two retrospective cohort studies of elite male professional football players drawn from the FIFA World Cup (national teams) and the UEFA European Cup (club teams). The World Cup cohort included players listed on official tournament rosters from 1930 to 1990. The European Cup cohort included players who appeared in quarterfinal, semifinal, or final matches from the 1955–56 to 1990–91 seasons. These periods were selected to maximize follow-up while ensuring reliable historical documentation of participation and vital status.

World Cup data were compiled from Wikidata rosters derived from FIFA documentation^16^. Roster positions were mapped to analytic positions (appendix p 2); center-backs and center-forwards were identified using an archival procedure drawing on Wikidata and corroborated with Transfermarkt’s historical sources^17^. Dates of birth and death were obtained from Wikidata using a prespecified matching protocol (appendix pp 3–6). After excluding 104 players (2.0%) with unresolvable vital status information and collapsing repeated appearances, the analytic cohort included 4,223 unique players. European Cup data were compiled from official UEFA competition records for players who appeared in any quarterfinal, semifinal, or final matches^18^. Dates of birth and death were obtained via Wikidata (appendix pp 3–6), and positions were coded using the same harmonized framework. After exclusions (53 players, 1.9%) and collapsing duplicates, the analytic cohort included 2,710 players.

### Variables

The primary exposure was playing position, harmonized into six analytic categories (Figure 1). This classification was applied consistently across the two studies and historical formations (appendix p 2). For example, center-back was also assigned to “stoppers” and “sweepers”, and “strikers” were classified as center-forward.

Models adjusted for region of origin, defined by grouping countries of birth into historically meaningful macro-regions to account for baseline mortality heterogeneity and differences in football environments and strategies. These are Northwestern Europe (plus North America), Southern Europe, and Central/Eastern Europe, sub-Saharan Africa, Asia, and the Maghreb/Middle East. The latter three regions represent very small sample sizes. They were retained in all adjusted models but interpreted cautiously.

Models were stratified by birth cohort rather than adjusted for birth year as a covariate. Stratification allows baseline hazards to vary flexibly across cohorts, accommodating cohort-specific mortality regimes while preserving estimation of covariate effects within comparable historical contexts.

### Statistical analysis

Survival differences across positions were first assessed using log-rank tests stratified by birth cohort. Let *t* denote survival time and *P* playing position. The stratified log-rank test evaluates whether the survivor functions differ across positions while allowing the baseline hazard to vary freely across birth cohort strata *s*. The null hypothesis is tested through:

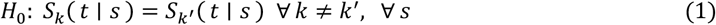

where *S*_*k*_(*t* ∣ *s*) is the survival function for position *k* within birth cohort stratum *s*. Rejection of the hypothesis indicates that survival distributions differ across playing positions after accounting for cohort-specific mortality patterns.

Primary analyses then estimated position-specific hazard ratios using Cox proportional hazards models, with midfielders specified as the reference given their intermediate physical demands. All Cox models were stratified by birth cohort to allow cohort-specific baseline hazards and adjusted for origin region. Survival curves by position were derived from adjusted Cox models and plotted as model-based survival functions. The mortality hazard for individual *i* at age *t*, stratified by birth cohort *s*, is:

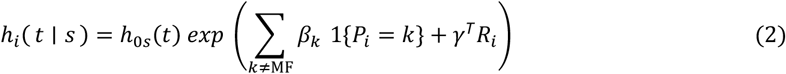

where *h*_0*s*_(*t*) is the baseline hazard specific to birth cohort *s, P*_*i*_ denotes position, *R*_*i*_ is a vector of origin region indicators, and *β*_*k*_ estimates the log hazard ratio for position *k* relative to midfielders. We thus estimate the ratio of the instantaneous all-cause mortality hazard – *exp* (*β*_*k*_) – at age *t* for players in position *k* relative to midfielders, adjusted for origin region and stratified by birth cohort.

To examine regional heterogeneity, we fitted interaction models between position and region within the stratified Cox framework:

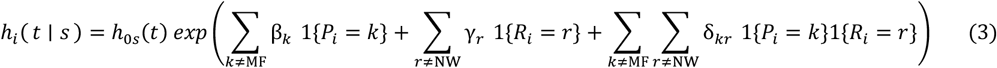

From these models, we computed adjusted marginal hazard ratios for each position-region combination by exponentiating the model linear predictor, yielding hazard ratio (*HR*) estimates expressed relative to the reference combination of midfielders from Northwestern Europe:

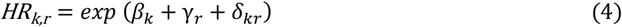

Robust variance estimators were used. All statistical tests were two-sided, with a significance threshold of *p* < 0.05.

## Results

### Position mortality risk

The World Cup cohort included 4,223 players, among whom 2,333 deaths were observed. The European Cup cohort comprised 2,710 players, with 1,126 observed deaths. Appendix pp 13–14 summarizes cohort composition by birth cohort, position, and region of origin. The World Cup and European Cup cohorts contained similar proportions of center-backs (12.6% and 13.0%, respectively) and center-forwards (11.0% and 11.7%, respectively). Crude mortality rates varied by position (appendix p 15) and across age-position strata, as shown in life tables (appendix p 16).

As shown in Table 1, stratified log-rank tests indicated statistically significant differences in survival across positions in both cohorts (World Cup: χ^2^=47.06; p<0.001; European Cup: χ^2^=11.74; p=0.039). In the World Cup cohort, goalkeepers experienced substantially fewer observed deaths than expected under the null hypothesis of equal survival, whereas center-backs and center-forwards exhibited excess observed mortality. A similar pattern was observed in the European Cup cohort, with excess observed deaths among center-backs and fewer than expected deaths among midfielders and non-central forwards. Because these comparisons are unadjusted and descriptive, we next estimated multivariable survival models to quantify position-specific mortality risks while accounting for cohort and regional differences.

**Table 1.** Log-rank tests for playing positions, stratified by birth cohort.

| Position | World Cup (1930–1990)<br>N = 4,223 |  |  | European Cup (1956–1991)<br>N = 2,710 |  |  |
| --- | --- | --- | --- | --- | --- | --- |
|  | observed<br>deaths | expected<br>deaths | %<br>difference<br>from<br>expected | observed<br>deaths | expected<br>deaths | %<br>difference<br>from<br>expected |
| Goalkeeper | 273 | 362.9 | -32.9% | 109 | 108.9 | 0.1% |
| Center-back | 294 | 256.4 | 12.8% | 175 | 148.3 | 15.3% |
| Defender (other) | 301 | 306.8 | -1.9% | 202 | 187.0 | 7.4% |
| Midfielder | 609 | 595.1 | 2.3% | 310 | 342.6 | -10.5% |
| Center-forward | 245 | 188.1 | 23.2% | 141 | 131.1 | 7.1% |
| Forward (other) | 611 | 623.8 | -2.1% | 189 | 208.2 | -10.1% |
|  | 2,333 |  |  | 1,126 |  |  |

*Notes.* Expected deaths are based on the null hypothesis of equal survival across playing positions within birth cohort strata. World Cup: Wald $\chi^2$ (5) = 47.01 (p<0.001). European Cup: Wald $\chi^2$ (5) = 11.74 (p=0.039).

### Adjusted mortality risk by position

Table 2 presents hazard ratios from Cox proportional hazards models stratified by birth cohort, and adjusted for region of origin (full results in appendices p 20 and p 21). In the World Cup cohort, goalkeepers had a significantly lower mortality hazard than midfielders (hazard ratio [HR], 0.73; 95% CI, 0.63–0.84; p<0.001), whereas center-forwards experienced 27% higher mortality hazard than midfielders during follow-up (HR, 1.27; 95% CI, 1.08–1.50; p=0.005). Mortality differences for center-backs were elevated but did not reach conventional levels of statistical significance in two-sided tests (HR, 1.13; 95% CI, 0.98–1.31; p=0.095). No significant differences were observed for other defenders or non-central forwards.

**Table 2.** Adjusted hazard ratios for mortality by position in World Cup and European Cup cohorts.

| Position | World Cup (1930–1990) |  | European Cup (1956–1991) |  |
| --- | --- | --- | --- | --- |
|  | Hazard ratio<br>(95% CI) | P value | Hazard ratio<br>(95% CI) | P value |
| Midfielder (reference) |  |  |  |  |
| Goalkeeper | 0.73 (0.63–0.84) | <0.001 | 1.11 (0.88–1.39) | 0.389 |
| Center-back | 1.13 (0.98–1.31) | 0.095 | 1.28 (1.07–1.55) | 0.009 |
| Defender (other) | 0.97 (0.84–1.11) | 0.613 | 1.20 (1.02–1.42) | 0.032 |
| Center-forward | 1.27 (1.08–1.50) | 0.005 | 1.19 (0.97–1.46) | 0.098 |
| Forward (other) | 0.95 (0.85–1.06) | 0.350 | 0.99 (0.82–1.19) | 0.878 |

*Notes.* Models were adjusted for region of origin and stratified by birth cohort, with robust standard errors and two-sided significance tests. Full model results are reported in appendix p 20. World Cup cohort: 4 223 subjects, 2 333 deaths, time at risk = 301,828, Wald $\chi^2$ (11) = 82.28 ( $p<0.001$ ). European Cup cohort: 2 710 subjects, 1 126 deaths, time at risk = 194,910, Wald $\chi^2$ (8) = 30.75 ( $p<0.001$ ).

In the European Cup cohort, center-backs experienced a significantly higher mortality hazard compared with midfielders (HR, 1.28; 95% CI, 1.07–1.55; p=0.009), as did other defenders (HR, 1.20; 95% CI, 1.02–1.42; p=0.032). Mortality hazards for center-forwards were elevated but not statistically significant (HR, 1.19; 95% CI, 0.97–1.46; p=0.098), while no significant differences were observed for goalkeepers or non-central forwards.

When positions were grouped at the level of positional lines (goalkeeper, defender, midfielder, forward), survival curves indicated higher mortality hazards for all outfield positions relative to goalkeepers, with limited differentiation among outfield positions (appendix p 22). In contrast, disaggregation into detailed positional roles revealed pronounced heterogeneity in survival. Using detailed position as exposure, Figure 2 displays adjusted model-based survival curves derived from Cox models controlling for birth cohort and region of origin. In the World Cup cohort (panel A), center-forwards exhibited the lowest survival probabilities across much of the age range, whereas in the European Cup cohort (panel B), center-backs and other defenders showed markedly lower survival. Consistent with the hazard ratio estimates, goalkeepers, midfielders, and other forwards (e.g., wingers, second strikers) exhibited comparatively higher survival, indicating that mortality differences were concentrated in the central defensive and attacking roles rather than uniformly distributed across positional lines.

**Figure 2.**
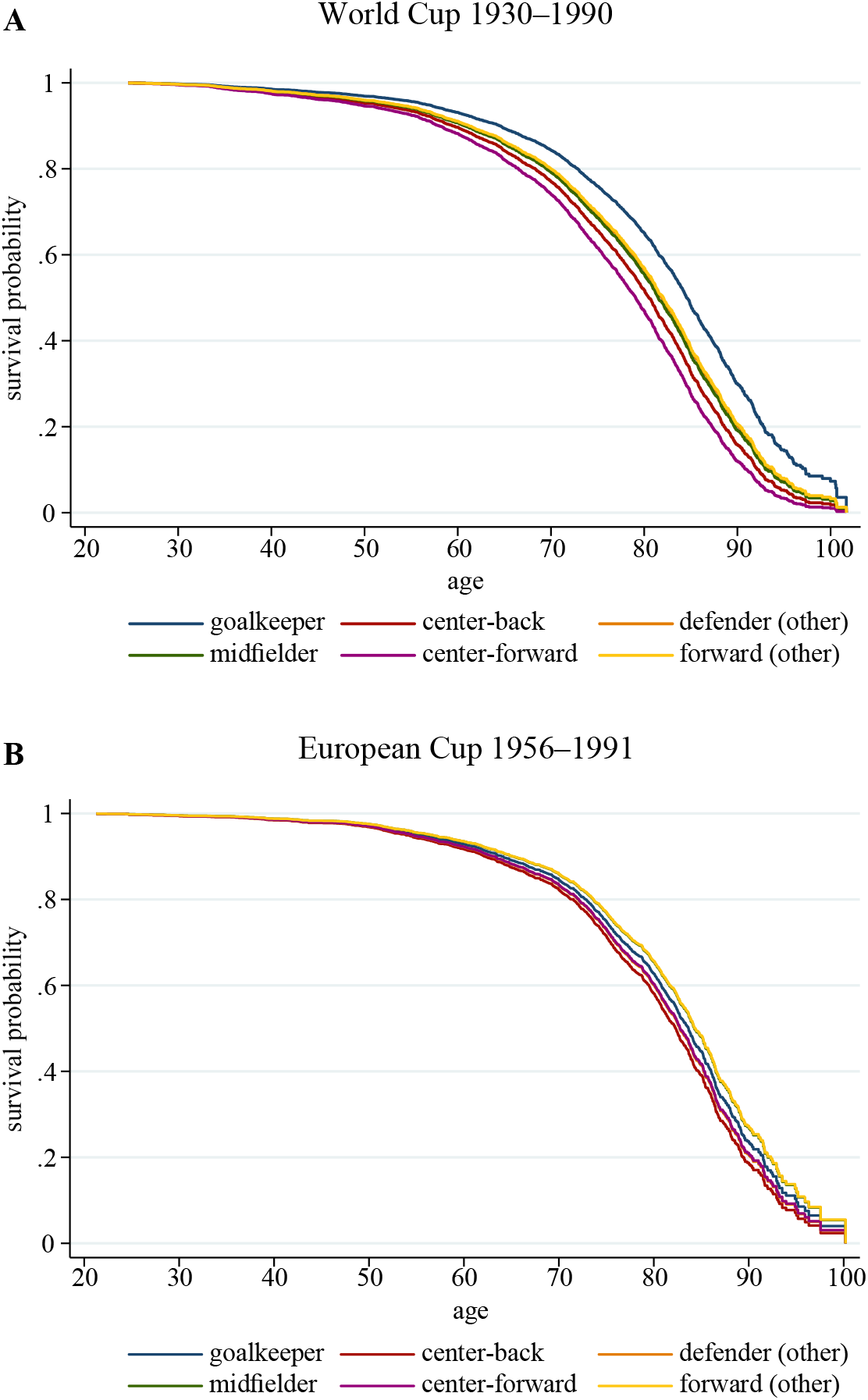
Adjusted survival curves by playing position. *Notes*. Model-based survival curves were derived from Cox proportional hazards models stratified by birth cohort and adjusted for region of origin, with age as the time scale. Curves are shown for the six analytic playing positions in the World Cup (A) and European Cup (B) cohorts. Corresponding survival curves using broader positional lines (goalkeeper, defender, midfielder, forward) for the World Cup cohort are shown in appendix p 22.

Restricted mean survival time (RMST) analyses provide complementary, absolute-scale interpretations of these differences (appendix p 23). In the World Cup cohort, center-forwards experienced the greatest loss of expected survival time relative to midfielders across the adult life course. By age 80, center-forwards had lost approximately 1 year and 5 months of life expectancy compared with midfielders, with smaller but progressively widening differences already evident by age 70 (8 months) and age 75 (1 year). Center-backs also exhibited reduced survival relative to midfielders, amounting to 7 months by age 80, whereas differences for other defenders and non-central forwards were minimal.

In the European Cup cohort, absolute differences in RMST were smaller, but displayed a similar ordering. Center-backs experienced the largest reduction in survival time relative to midfielders, amounting to 1 year by age 80, with deficits emerging gradually from midlife onward. Other positions showed more modest losses, yet goalkeepers and non-central forwards exhibited survival profiles closely aligned with midfielders. Together, the RMST estimates illustrate that excess mortality among central defensive and attacking roles translates into meaningful differences in expected years of life, particularly at older ages.

### Region heterogeneity

Figure 3 presents adjusted marginal hazard ratios from Cox models including interactions between playing position and region of origin, stratified by birth cohort and adjusted for all main effects. These margins summarize the combined position-specific and regional associations relative to midfielders from Northwestern Europe and are therefore informative even when individual interaction terms do not reach conventional levels of statistical significance. Full interaction estimates, as well as estimates for regions with limited representation (Asia, sub-Saharan Africa, Maghreb/Middle East), are reported in appendices pp 24–25 and pp 26–27. Across both cohorts, substantial heterogeneity was observed in how position-related mortality risk varied by region.

**Figure 3.**
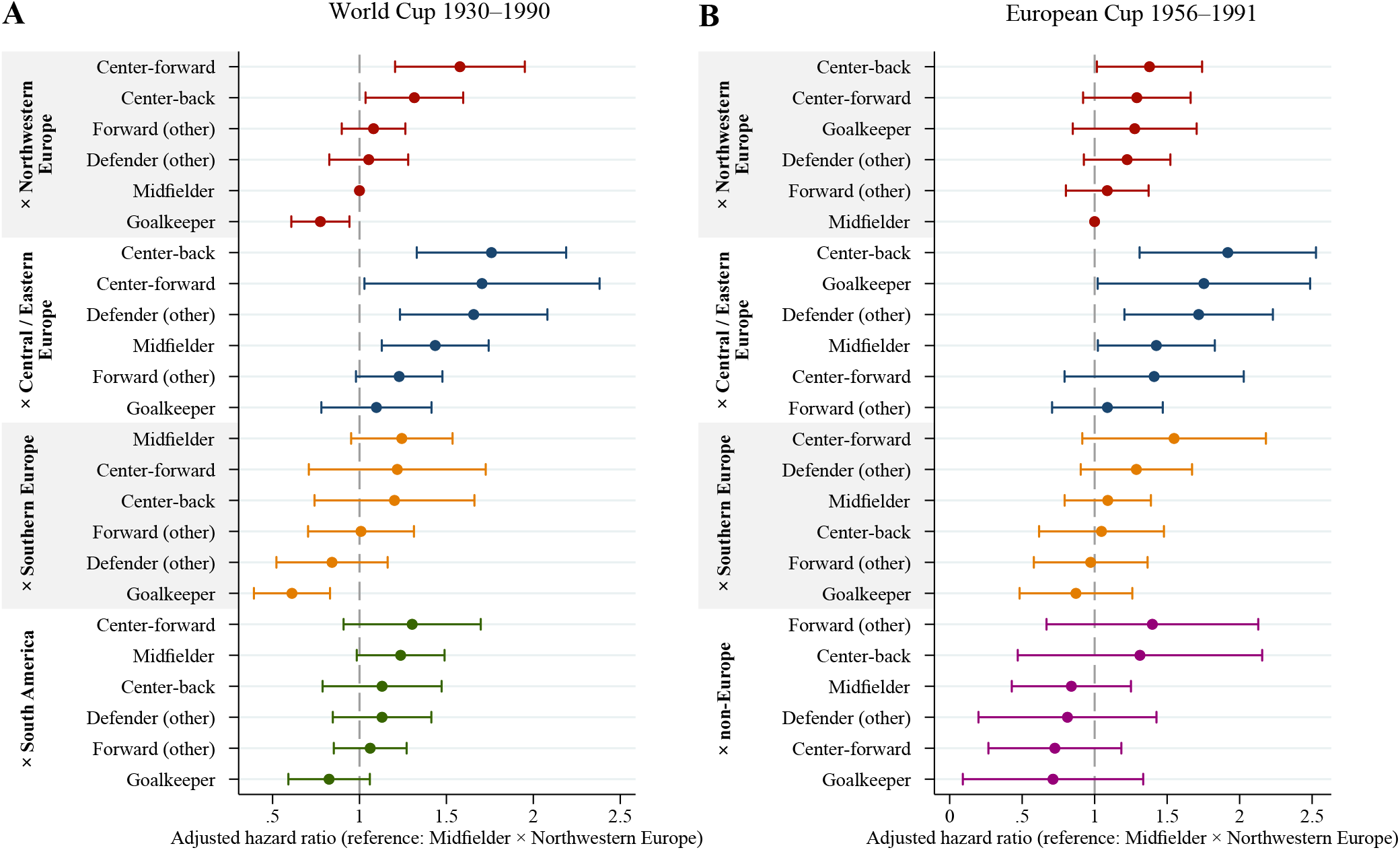
Adjusted marginal hazard ratios for mortality by playing position and region of origin. *Notes*. The figure displays adjusted marginal hazard ratios derived from Cox proportional hazards models including interactions between playing position and region of origin, stratified by birth cohort and adjusted for all main effects. Estimates were obtained using model-based marginal predictions on the hazard ratio scale. Results are shown for both the World Cup (A) and European Cup (B) cohorts. Full interaction model estimates are reported in appendix pp 24–25. All marginal estimates, including for the regions with limited representation, are reported in appendix pp 26–27.

In the World Cup cohort, elevated mortality risks for central playing roles were most pronounced among players from Northwestern Europe and Central/Eastern Europe (Figure 3, panel A). Among Northwestern European players, center-forwards exhibited the highest adjusted mortality hazard (HR, 1.58; 95% CI, 1.20–1.95), followed closely by center-backs (HR, 1.32; 95% CI, 1.03–1.60), relative to midfielders from the same region. Corresponding mortality risks were even higher among Central/Eastern European players, with elevated hazards for center-backs (HR, 1.76; 95% CI, 1.33–2.19) and center-forwards (HR, 1.70; 95% CI, 1.03–2.38). In contrast, goalkeepers experienced a clear survival advantage in Northwestern Europe (HR, 0.78; 95% CI, 0.61–0.94), whereas this protective association was attenuated in Central/Eastern Europe (HR, 1.10; 95% CI, 0.78–1.41), where goalkeeper mortality approximated that of outfield players.

Comparable patterns were observed in the European Cup cohort (Figure 3, panel B). Among players from Central/Eastern Europe, center-backs exhibited an estimated 92% higher mortality hazard than Northwestern European midfielders (HR, 1.92; 95% CI, 1.31–2.53), with similarly elevated risks observed for other defenders (HR, 1.72; 95% CI, 1.21–2.23), relative to Northwestern European midfielders. Northwestern European center-backs remained at elevated risk (HR, 1.38; 95% CI, 1.02–1.74), whereas positional differences were less pronounced in players from Southern Europe and non-European regions (e.g., South America). As in the World Cup cohort, goalkeepers showed relatively favorable survival.

Although some individual position-by-region estimates were based on small numbers and were therefore associated with wider confidence intervals, the consistency of these marginal patterns suggests that excess mortality associated with central defensive and attacking roles is concentrated within specific regional contexts rather than uniformly distributed across elite football populations.

## Discussion

In this retrospective analysis of elite male football players active across much of the 20th century, long-term all-cause mortality differed systematically by playing position. Across two independent cohorts drawn from international and club competition, central attacking and defensive roles – particularly center-forwards and, in most contexts, center-backs – were associated with higher mortality risk relative to midfielders, whereas goalkeepers generally experienced more favorable survival. These patterns were most pronounced in regions where direct styles of play and aerial contests historically predominated, underscoring the importance of positional role and tactical context in shaping long-term mortality outcomes.

Notably, center-backs did not consistently exhibit elevated mortality risk in the World Cup cohort after multivariable adjustment, despite doing so in the European Cup cohort. One plausible explanation is that earlier birth cohorts in the World Cup data played in eras when defensive responsibilities – and exposure to aerial play – were more diffusely distributed across defensive positions rather than concentrated in specialized central defenders. In contrast, the center-forward role has long functioned as a focal point for aerial contests, including challenges for long balls, goal kicks, and crosses, implying sustained exposure to high-impact play across tactical eras.

These findings contribute to a growing literature linking repetitive head impacts in contact sports to long-term mortality. For example, in football, former National Football League players have been shown to experience elevated mortality risk, with heterogeneity by playing position^19^. Our results provide population-level evidence that similar hypothesized exposure patterns in elite football may also translate into differential mortality risk. Moreover, by demonstrating that such heterogeneity was evident among elite players competing throughout the 20th century, further specified by recognizable regional playing styles, this study situates contemporary concerns within a longer historical perspective. Unlike prior work focused on single national leagues or later cohorts, the present analysis draws on international and club tournament data spanning multiple decades and regions, strengthening the generalizability of observed patterns.

Several limitations merit consideration. Playing position served as a proxy for cumulative exposure to head impacts and physical contact, as direct measures of heading frequency, collision severity, or concussion history were unavailable, particularly for early eras. Mortality outcomes were ascertained from archival sources rather than administrative registries, and cause-specific mortality could not be examined. Estimates for some position-region combinations were based on small numbers, and residual confounding by unmeasured factors cannot be excluded; accordingly, associations should not be interpreted as causal effects.

Despite these limitations, the findings have implications for research on elite athletes and for ongoing debates surrounding football-related head impacts. The concentration of elevated mortality risk in central playing roles suggests that long-term health risks may be shaped not simply by participation in football *per se*, but by sustained exposure to specific tactical demands over the life course. This has relevance for contemporary monitoring of player health, for the evaluation of position-specific exposure pathways^11^, and for the design of preventive strategies that account for heterogeneity within the sport. More broadly, the results highlight the value of historically grounded, cross-national analyses for understanding how sport-specific exposures accumulate over time and translate into later-life health outcomes.

## Data Availability

The data used in this study were derived from publicly accessible archival sources, including Wikidata, official FIFA World Cup documentation, UEFA historical records, and Transfermarkt. The analytic dataset and statistical code generated for this study are available from the corresponding author upon reasonable request.

## Data availability statement

Data used in this study were derived from publicly accessible archival sources. The analytic datasets and code for statistical analysis are available from the corresponding author on reasonable request.

## Contributions

Conceptualisation: Humphreys, Witteveen

Methodology: Witteveen

Data curation: Witteveen

Formal analysis: Witteveen

Investigation: Witteveen

Resources: Witteveen

Software: Witteveen

Visualisation: Witteveen

Validation: Witteveen

Supervision: Humphreys

Project administration: Witteveen

Funding acquisition: Not applicable

Writing – original draft: Witteveen

Writing – review & editing: Humphreys, Witteveen

