## Supplement for "Playing position and long-term mortality among elite male football players, 1930–1990"

Dirk Witteveen, PhD[1,2]

David K. Humphreys, PhD[1]

[1] Department of Social Policy and Intervention, University of Oxford, Oxford, UK

Version: February 16, 2026

Appendix contents:

Appendix 1: Historical positions and analytic position classification. (p 2)

Appendix 2: Methodological considerations. (pp 3 – 12)

Appendix 3: Cohort composition by birth cohort, position, and region of origin. (p 13 – 14)

Appendix 4: Mortality rates by position. (p 15)

Appendix 5: Life tables stratified by position. (pp 16 – 19)

Appendix 6: Cox regressions estimates. (p 20)

Appendix 7: Cox regressions estimates with alternative reference categories. (p 21)

Appendix 8: Position-line-specific survival (World Cup cohort). (p 22)

Appendix 9: Restricted mean survival times (RMST). (p 23)

Appendix 10: Cox regression interaction model estimates. (pp 24 – 25)

Appendix 11: Marginal hazard ratios from Cox regression interaction model estimates. (pp 26 – 27)

**Appendix 1**. Historical positions and analytic position classification.

| Historical position | Analytic position |
|---|---|
| Goalkeeper | Goalkeeper |
| Full-back | Defender (other) |
| Wing-back | Defender (other) |
| Left-back | Defender (other) |
| Right-back | Defender (other) |
| Defender | Defender (other) |
| Center-back | Center-back |
| Central defender | Center-back |
| Stopper | Center-back |
| Center-half | Center-back |
| Libero (defensive) | Center-back |
| Midfielder | Midfielder |
| Attacking midfielder | Midfielder |
| Left-half | Midfielder |
| Right-half | Midfielder |
| Left midfield | Midfielder |
| Right midfield | Midfielder |
| Half-back | Midfielder |
| Winger | Forward (other) |
| Outside forward | Forward (other) |
| Inside forward | Forward (other) |
| Second striker | Forward (other) |
| Center-forward | Center-forward |
| Striker | Center-forward |

**Appendix 2**. Methodological considerations.

*Data provenance and archival sources*

Player-level data for both cohorts were derived from publicly accessible archival sources that compile historical competition records for elite professional football. For the World Cup cohort, Wikidata served as a structured aggregation layer for official FIFA national squad lists, drawing on FIFA tournament documentation, national federation publications, and established historical compilations. For the European Cup cohort, official UEFA competition records provided match appearance data for advanced-stage fixtures. Wikidata was additionally used to harmonize biographical information across cohorts and to ascertain vital status.

Because no centralized administrative registry exists for elite professional football players active during the mid-20th century, the use of community-curated archival databases represents the most comprehensive and transparent approach to reconstructing long-term cohorts with sufficient follow-up for mortality analyses. All data extraction and linkage procedures were automated where possible and supplemented by targeted manual verification in ambiguous cases.

Data extraction, linkage, and cohort construction were automated using custom Python scripts, with statistical analyses conducted in Stata. Automation was used to ensure reproducibility of the initial dataset and consistent application of inclusion, exclusion, and linkage rules across cohorts, while targeted manual verification was applied in ambiguous cases.

*Data extraction procedures and normalization*

**World Cup cohort**. Tournament rosters were obtained from Wikidata records titled "[Year] FIFA World Cup squads," which compile official national squad lists derived from FIFA historical tournament documentation and related archival sources. A custom Python pipeline parsed roster tables for each participating country and extracted player name, nationality, date of birth, and roster-listed position. Text fields were normalized to reduce duplication arising from diacritics, alternative transliterations, and punctuation. Dates of birth were standardized to ISO-8601 format when available. When birth dates were missing or ambiguously formatted, partial or inconsistent entries were conservatively coded as missing and reviewed.

**European Cup cohort**. Match appearance data were extracted from publicly accessible UEFA competition records for players who appeared on the field in quarterfinal, semifinal, or final matches between the 1955–1956 and 1990–1991 seasons. Match appearance data were extracted using custom Python scripts from publicly accessible UEFA competition records and included player name, club, season, and match stage. UEFA records did not consistently include dates of birth; therefore, dates of birth were obtained via linkage to Wikidata and standardized to ISO-8601 format.

*Mortality linkage and validation*

Vital status was ascertained through linkage to the Wikidata API. Candidate entities were retrieved based on full-name matching and restricted to entities classified as *human* and having the occupation "association football player." Where multiple candidate entities were identified, records were flagged for manual review by the investigators. Linkage preference was given to entities whose recorded date (or year) of birth matched the value extracted from competition records. Additional corroborating metadata (e.g., nationality, club affiliations, or tournament participation) were used in manual review where available.

Dates of death were extracted when present and standardized to ISO-8601 format. When only the year of death was recorded, the death date was retained as year-only and assigned July 1 for survival-time calculations to avoid systematic bias toward early- or late-year deaths (12 times across both cohorts combined). Players for whom no reliable evidence of death or survival status could be established after archival review were excluded from the analytic cohorts.

For the World Cup cohort, the initial dataset comprised 5,222 player-tournament observations. After excluding 104 players (2.0%) with unresolvable or conflicting vital status information and collapsing repeated tournament appearances, the final analytic cohort included 4,223 unique players. For the European Cup cohort, after collapsing duplicates across rounds and seasons and conducting additional archival checks (see below), 53 players (1.9%) were excluded due to unverifiable vital status, yielding a final analytic cohort of 2,710 unique players.

*Playing position coding and historical harmonization*

Playing positions were harmonized using a standardized classification scheme designed to accommodate historical variation in football tactics and position terminology. Original position labels were first mapped to four primary positional lines: goalkeeper, defender, midfielder, and forward.

Additional sub-positional distinctions for center-back and center-forward were assigned only when supported by consistent career-level descriptions across multiple archival sources. Sub-positional coding relied primarily on Wikidata and Transfermarkt and was supplemented by corroborating historical materials (e.g., contemporary match reports and player biographies) when needed. Coding explicitly considered historically specific terminology – e.g., stopper, sweeper, libero, half-back, inside forward, center-half – and prevailing formations of the era.

Transfermarkt is widely used by journalists, scouts, analysts, and technical staff within professional football as a reference source for player career histories and positional information. Its coverage and crowd-curated structure have been examined in journalistic reporting on collective intelligence in sports data environments[22]. In addition, Transfermarkt data have been used in peer-reviewed research on player valuation and labor-market returns in professional football, including studies of the economic payoffs to player characteristics and technical skills[23]. In the present study, Transfermarkt was used exclusively to corroborate career-level positional descriptions and was not relied upon for outcome ascertainment.

Central defensive and attacking roles were identified conservatively to minimize misclassification. When sources unanimously placed a player within a positional line but did not unambiguously support a central sub-role, the player was retained in the broader category (defender [other] or forward [other]). Cases with conflicting or insufficient evidence were left unclassified at the sub-positional level. The full mapping of historical position labels to analytic categories is provided in appendix 1 p 2. When sources did not consistently support a central role, players were retained in their broader positional category.

*Handling of repeated observations and deduplication*

**World Cup cohort.** Because players could appear on tournament rosters across multiple World Cup editions, player-tournament observations were collapsed to unique individuals after mortality linkage. Each player contributed a single survival record.

**European Cup cohort.** Players could appear in multiple rounds within a season and across multiple seasons, potentially with different clubs. Therefore, duplicate records were collapsed after aggregation across rounds and seasons such that each player contributed a single survival record.

As part of data quality assurance, records were flagged for review by the researchers when birth dates or death dates implied implausible ages at match appearance or death (e.g., appearances at extreme ages inconsistent with professional play), and these cases were resolved via archival checks where possible.

*Birth cohort stratification*

Rather than adjusting for birth year as a continuous covariate, analyses were stratified by birth cohort to allow the baseline hazard to vary flexibly across historically distinct mortality regimes. Cohort definitions were tailored to the historical coverage of each dataset to reflect discontinuities in early-life conditions and secular changes in professionalization and match intensity, preserving estimation of covariate associations within comparable historical contexts. For the World Cup cohort, birth cohorts were defined to align with major geopolitical and epidemiological eras characterized by discontinuities in early-life conditions (pre-World War I, World War I, interwar period, World War II, and postwar). For the European Cup cohort, which spans a narrower and more continuous range of birth years, approximately decadal cohorts were used to capture generational differences in professionalization and match intensity.

*Region of origin classification*

Region of origin was defined using players' countries of birth and grouped into historically meaningful macro-regions to capture broad differences in baseline mortality risk, early-life conditions, and football development environments. European regions were subdivided to preserve heterogeneity, while regions with limited representation were grouped more coarsely to maintain analytic stability. Regions with small sample sizes were retained in adjusted models but interpreted cautiously.

**Table S2.** Region labels.

| | World Cup (1930–1990) N = 4 223 | | European Cup (1956–1991) N = 2 710 | |
|---|---|---|---|---|
| | Region | Observations | Region | Observations |
| Albania | Central / Eastern Europe | 0 | Central / Eastern Europe | 1 |
| Algeria | Maghreb / Middle East | 34 | non-Europe | 1 |
| Angola | Sub-Saharan Africa | 0 | non-Europe | 3 |
| Argentina | South / Central America | 171 | non-Europe | 21 |
| Armenia | Central / Eastern Europe | 0 | Central / Eastern Europe | 4 |
| Australia | Northwestern Europe | 22 | non-Europe | 2 |
| Austria | Northwestern Europe | 110 | Northwestern Europe | 82 |
| Belarus | Central / Eastern Europe | 0 | Central / Eastern Europe | 5 |
| Belgium | Northwestern Europe | 141 | Northwestern Europe | 94 |
| Bolivia | South / Central America | 18 | non-Europe | 0 |
| Brazil | South / Central America | 220 | non-Europe | 30 |
| Bulgaria | Central / Eastern Europe | 84 | Central / Eastern Europe | 71 |
| Cameroon | Sub-Saharan Africa | 40 | non-Europe | 0 |
| Canada | Northwestern Europe | 22 | non-Europe | 0 |
| Chile | South / Central America | 113 | non-Europe | 0 |
| Colombia | South / Central America | 44 | non-Europe | 0 |
| Costa Rica | South / Central America | 22 | non-Europe | 0 |
| Croatia | Central / Eastern Europe | 0 | Central / Eastern Europe | 10 |
| Cuba | South / Central America | 7 | non-Europe | 0 |
| Czechia | Central / Eastern Europe | 0 | Central / Eastern Europe | 44 |
| Czechoslovakia | Central / Eastern Europe | 150 | Central / Eastern Europe | 36 |
| DR Congo (Zaire) | Sub-Saharan Africa | 22 | non-Europe | 3 |
| Denmark | Northwestern Europe | 22 | Northwestern Europe | 50 |
| Dutch East Indies | Asia | 9 | non-Europe | 0 |
| East Germany | Central / Eastern Europe | 22 | Northwestern Europe | 43 |
| Egypt | Maghreb / Middle East | 34 | non-Europe | 0 |
| El Salvador | South / Central America | 42 | non-Europe | 0 |
| England | Northwestern Europe | 145 | Northwestern Europe | 149 |
| Finland | Northwestern Europe | 0 | Northwestern Europe | 12 |
| France | Northwestern Europe | 152 | non-Europe | 108 |
| Germany | Northwestern Europe | 33 | Northwestern Europe | 279 |
| Ghana | Sub-Saharan Africa | 0 | non-Europe | 1 |
| Greece | Southern Europe | 0 | Southern Europe | 44 |
| Haiti | South / Central America | 22 | non-Europe | 0 |
| Honduras | South / Central America | 22 | non-Europe | 0 |
| Hungary | Central / Eastern Europe | 148 | Central / Eastern Europe | 98 |
| Iceland | Northwestern Europe | 0 | Northwestern Europe | 3 |
| Italy | Southern Europe | 196 | Southern Europe | 189 |
| Iran | Maghreb / Middle East | 22 | non-Europe | 0 |
| Iraq | Maghreb / Middle East | 22 | non-Europe | 0 |
| Ireland (Republic) | Northwestern Europe | 22 | Northwestern Europe | 15 |

| | | | | |
|---|---|---|---|---|
| Israel | Northwestern Europe | 22 | Northwestern Europe | 0 |
| Ivory Coast | Sub-Saharan Africa | 0 | non-Europe | 1 |
| Japan | Asia | 0 | non-Europe | 1 |
| Liberia | Sub-Saharan Africa | 0 | non-Europe | 1 |
| Kuwait | Maghreb / Middle East | 22 | non-Europe | 0 |
| Luxembourg | Northwestern Europe | 0 | Northwestern Europe | 2 |
| Mexico | South / Central America | 147 | non-Europe | 1 |
| Monaco | Northwestern Europe | 0 | Northwestern Europe | 1 |
| Montenegro | Central / Eastern Europe | 0 | Central / Eastern Europe | 1 |
| Morocco | Maghreb / Middle East | 40 | non-Europe | 2 |
| Netherlands | Northwestern Europe | 91 | Northwestern Europe | 143 |
| New Zealand | Northwestern Europe | 22 | non-Europe | 0 |
| Nigeria | Sub-Saharan Africa | 0 | non-Europe | 1 |
| North Korea | Asia | 8 | non-Europe | 0 |
| North Macedonia | Central / Eastern Europe | 0 | Central / Eastern Europe | 5 |
| Northern Ireland | Northwestern Europe | 55 | Northwestern Europe | 16 |
| Norway | Northwestern Europe | 22 | Northwestern Europe | 4 |
| Paraguay | South / Central America | 64 | non-Europe | 2 |
| Peru | South / Central America | 73 | non-Europe | 1 |
| Poland | Central / Eastern Europe | 83 | Central / Eastern Europe | 68 |
| Portugal | Southern Europe | 44 | Southern Europe | 102 |
| Romania | Central / Eastern Europe | 90 | Central / Eastern Europe | 57 |
| Russia | Central / Eastern Europe | 0 | Central / Eastern Europe | 18 |
| San Marino | Southern Europe | 0 | Southern Europe | 1 |
| Scotland | Northwestern Europe | 123 | Northwestern Europe | 181 |
| Senegal | Sub-Saharan Africa | 0 | non-Europe | 2 |
| Serbia | Central / Eastern Europe | 0 | Central / Eastern Europe | 143 |
| Slovakia | Central / Eastern Europe | 0 | Central / Eastern Europe | 21 |
| Slovenia | Central / Eastern Europe | 0 | Central / Eastern Europe | 1 |
| South Africa | Sub-Saharan Africa | 0 | non-Europe | 2 |
| South Korea | Asia | 54 | non-Europe | 0 |
| Soviet Union | Central / Eastern Europe | 111 | Central / Eastern Europe | 75 |
| Spain | Southern Europe | 151 | Southern Europe | 214 |
| Sweden | Northwestern Europe | 148 | Northwestern Europe | 87 |
| Switzerland | Northwestern Europe | 104 | Northwestern Europe | 73 |
| Tunesia | Maghreb / Middle East | 22 | non-Europe | 0 |
| Turkey | Maghreb / Middle East | 22 | Southern Europe | 47 |
| Ukraine | Central / Eastern Europe | 0 | Central / Eastern Europe | 21 |
| United A. Emirates | Maghreb / Middle East | 22 | non-Europe | 0 |
| United States | Northwestern Europe | 70 | non-Europe | 0 |
| Uruguay | South / Central America | 161 | non-Europe | 1 |
| Wales | Northwestern Europe | 22 | Northwestern Europe | 14 |
| West Germany | Northwestern Europe | 152 | Northwestern Europe | 0 |
| Yugoslavia | Central / Eastern Europe | 145 | Central / Eastern Europe | 1 |
| Zimbabwe | Sub-Saharan Africa | 0 | non-Europe | 1 |

*Statistical analyses and robustness*

Survival analyses were conducted using stratified Cox proportional hazards models with age as the time scale and birth cohort as the stratification variable. Models were adjusted for region of origin, and robust variance estimators were used to account for mild clustering and potential departures from model assumptions. Analyses were conducted separately for the World Cup and European Cup cohorts.

Proportional hazards assumptions were evaluated using Schoenfeld residual tests and visual inspection of log-log survival plots. In the World Cup cohort, Schoenfeld residual tests indicated departures from proportional hazards for the center-forward position and for selected region indicators. Accordingly, sensitivity analyses allowing time-varying coefficients for these covariates were conducted; these analyses indicated that position-specific mortality differences varied over age but did *not* alter the direction or substantive interpretation of the primary results. In contrast, no meaningful global departures from proportional hazards were detected in the European Cup cohort, and standard stratified Cox models were retained.

Interaction models between playing position and region of origin were estimated within the same stratified Cox framework to examine regional heterogeneity in position-specific mortality risk. Region-specific position estimates were summarized using model-based marginal hazard ratios.

*Alternate specification of the main model: Table 2 (Appendix 7).*

We chose the outfield position midfielder as the reference category in the main models (Table 2) given their intermediate physical demands. Appendix 7 p 21 shows specifications with alternative reference categories. Inclusion and exclusion of players from United States, Canada, and Israel in the Northwestern Europe (“plus”) region does not alter the estimates (therefore not shown).

*Restricted Mean Survival Time (RMST) procedure (Appendix 9).*

To complement hazard ratio-based analyses, we estimated restricted mean survival time (RMST) and derived years of life lost before specified age thresholds (appendix 9, p 23). RMST provides an absolute, time-scale-interpretable summary of survival that does not rely on proportional hazards assumptions and is particularly informative when differences in risk accumulate gradually over the life course. We estimated RMST using flexible parametric survival models (Royston-Parmar models) fitted on the age time scale. Both region and birth cohort were included as a categorical covariate to account for secular changes in mortality risk across cohorts. The baseline hazard was modeled using restricted cubic splines with five degrees of freedom, and robust standard errors were used throughout. For a given age threshold τ, restricted mean survival time was defined as the expected number of years lived up to that age:

$$RMST(\tau) = \int_{0}^{t} S(t)dt$$

where $S(t)$ denotes the estimated survival function at age $t$. RMST was estimated numerically by integrating the model-predicted survival curve over a fine age grid.

Years of life lost before age $\tau$were then calculated as:

$$YLL(\tau) = \tau - RMST(\tau)$$

To obtain marginal, population-representative estimates, we standardized RMST and years of life lost over the observed joint distribution of origin region and birth cohort. Specifically, for each playing position, survival functions were predicted for all individuals in the estimation sample under the counterfactual assumption that all individuals occupied that position while retaining their observed covariate values. Predicted survival probabilities were then averaged across individuals at each age, yielding standardized survival curves from which RMST and years of life lost were computed. Estimates are reported for age thresholds of 60, 65, 70, 75, and 80 years.

Several considerations are important when interpreting RMST-based estimates. First, RMST depends explicitly on the age thresholds $\tau$; differences between groups may appear modest at younger ages but grow at older thresholds as small: risk differentials accumulate *over time*. Thus, results are presented across multiple age cutoffs rather than a single threshold. Second, RMST estimation requires correct specification of the survival model over the entire age range up to $\tau$. Flexible parametric models mitigate this concern by allowing the baseline hazard to vary smoothly with age, but estimates at older ages may still be sensitive to sparse data. Third, unlike hazard ratios, RMST is a functional of the entire survival curve, not a single regression coefficient. As a result, RMST estimates are more computationally involved and more sensitive to modeling choices (e.g., standardization strategy). While hazard ratios summarize relative instantaneous risk and are comparatively straightforward to estimate and interpret, RMST provides an absolute, time-scale-based measure that is often more intuitive but requires greater care in implementation. Finally, as with all observational analyses, RMST and years-lost estimates reflect associations conditional on measured covariates and should *not* be interpreted as strictly causal effects.

*Replication*

Code for statistical analysis are available from the corresponding author on reasonable request.

**Appendix 3**. Cohort composition by birth cohort, position, and region of origin.

| World Cup (1930–1990) N = 4,223 | | |
|---|---|---|
| Birth cohort | | |
| 1892-1909 (pre-WWI) | 399 | 9.5% |
| 1910-1919 (WWI) | 366 | 8.7% |
| 1920-1929 (interwar) | 492 | 11.7% |
| 1930-1945 (Depression/WWII) | 1,057 | 25.0% |
| 1946-1970 (postwar) | 1,909 | 45.2% |
| Position | | |
| Goalkeeper | 523 | 12.4% |
| Center-back | 533 | 12.6% |
| Defender (other) | 657 | 15.6% |
| Midfielder | 1,193 | 28.3% |
| Center-forward | 465 | 11.0% |
| Forward (other) | 852 | 20.2% |
| Region | | |
| Northwestern Europe (and North America) | 1,500 | 35.5% |
| Central / Eastern Europe | 833 | 19.7% |
| Southern Europe | 391 | 9.3% |
| South / Central America | 1,126 | 26.7% |
| Maghreb / Middle East | 240 | 5.7% |
| Asia | 71 | 1.7% |
| Sub-Saharan Africa | 62 | 1.5% |

| European Cup (1956–1991)<br>N = 2,710 | | |
|---|---|---|
| Birth cohort | | |
| 1918-1929 | 166 | 6.1% |
| 1930-1939 | 590 | 21.8% |
| 1940-1949 | 717 | 26.5% |
| 1950-1959 | 762 | 28.1% |
| 1960-1972 | 475 | 17.5% |
| Position | | |
| Goalkeeper | 225 | 8.3% |
| Center-back | 353 | 13.0% |
| Defender (other) | 546 | 20.2% |
| Midfielder | 836 | 30.9% |
| Center-forward | 318 | 11.7% |
| Forward (other) | 432 | 15.9% |
| | 2,710 | |
| Region | | |
| Northwestern Europe (and North America) | 1,247 | 46.0% |
| Central / Eastern Europe | 680 | 25.1% |
| Southern Europe | 596 | 22.0% |
| Non-Europe | 187 | 6.9% |

**Appendix 4**. Mortality rates by position.

| | World Cup (1930–1990) | | | |
|---|---|---|---|---|
| Position | N | Deaths | Crude mortality rate (per 1 000 person-years) | Median attained age at exit |
| Goalkeeper | 523 | 273 | 7.1 | 73.9 |
| Center-back | 533 | 294 | 7.9 | 70.0 |
| Defender (other) | 657 | 301 | 6.4 | 71.9 |
| Midfielder | 1193 | 609 | 7.2 | 71.0 |
| Center-forward | 465 | 245 | 7.7 | 69.2 |
| Forward (other) | 852 | 611 | 9.7 | 74.0 |

| | European Cup (1956–1991) | | | |
|---|---|---|---|---|
| Position | N | Deaths | Crude mortality rate (per 1 000 person-years) | Median attained age at exit |
| Goalkeeper | 225 | 109 | 6.6 | 73.1 |
| Center-back | 353 | 175 | 6.9 | 72.4 |
| Defender (other) | 546 | 202 | 5.2 | 71.8 |
| Midfielder | 836 | 310 | 5.2 | 71.8 |
| Center-forward | 318 | 141 | 6.2 | 71.1 |
| Forward (other) | 432 | 189 | 6.0 | 74.3 |

**Appendix 5**: Life tables stratified by position.

| | | World Cup (1930–1990) N = 4 ,223 | | | | |
|---|---|---|---|---|---|---|
| Position | Interval | At risk | Deaths | Censored | Cum. survival prob. | Interval mortality risk |
| Goalkeeper | | | | | | |
| | 20 - 30 | 523 | 1 | 0 | 0.998 | 0.002 |
| | 30 - 40 | 522 | 6 | 0 | 0.987 | 0.013 |
| | 40 - 50 | 516 | 12 | 0 | 0.964 | 0.036 |
| | 50 - 60 | 504 | 21 | 11 | 0.923 | 0.077 |
| | 60 - 70 | 472 | 44 | 96 | 0.827 | 0.173 |
| | 70 - 80 | 332 | 73 | 95 | 0.615 | 0.385 |
| | 80 - 90 | 164 | 90 | 42 | 0.228 | 0.772 |
| | 90 - 100 | 32 | 24 | 6 | 0.039 | 0.961 |
| | > 100 | 2 | 2 | 0 | 0.000 | 1.000 |
| Center-back | | | | | | |
| | 20 - 30 | 533 | 1 | 0 | 0.998 | 0.002 |
| | 30 - 40 | 532 | 14 | 0 | 0.972 | 0.028 |
| | 40 - 50 | 518 | 16 | 0 | 0.942 | 0.058 |
| | 50 - 60 | 502 | 30 | 21 | 0.884 | 0.116 |
| | 60 - 70 | 451 | 66 | 119 | 0.735 | 0.265 |
| | 70 - 80 | 266 | 86 | 75 | 0.459 | 0.542 |
| | 80 - 90 | 105 | 73 | 22 | 0.102 | 0.898 |
| | 90 - 100 | 10 | 7 | 2 | 0.023 | 0.977 |
| | > 100 | 1 | 1 | 0 | 0.000 | 1.000 |
| Defender (other) | | | | | | |
| | 20 - 30 | 657 | 1 | 0 | 0.999 | 0.001 |
| | 30 - 40 | 656 | 12 | 0 | 0.980 | 0.020 |
| | 40 - 50 | 644 | 9 | 0 | 0.967 | 0.034 |
| | 50 - 60 | 635 | 26 | 16 | 0.926 | 0.074 |
| | 60 - 70 | 593 | 54 | 156 | 0.829 | 0.171 |
| | 70 - 80 | 383 | 91 | 137 | 0.589 | 0.411 |
| | 80 - 90 | 155 | 95 | 40 | 0.175 | 0.825 |
| | 90 - 100 | 20 | 13 | 7 | 0.037 | 0.963 |
| Midfielder | | | | | | |
| | 20 - 30 | 1193 | 5 | 0 | 0.996 | 0.004 |
| | 30 - 40 | 1188 | 18 | 0 | 0.981 | 0.019 |
| | 40 - 50 | 1170 | 28 | 0 | 0.957 | 0.043 |
| | 50 - 60 | 1142 | 67 | 48 | 0.900 | 0.100 |
| | 60 - 70 | 1027 | 114 | 278 | 0.784 | 0.216 |
| | 70 - 80 | 635 | 185 | 178 | 0.519 | 0.481 |
| | 80 - 90 | 272 | 164 | 75 | 0.156 | 0.844 |

| | | | | | | |
|---|---|---|---|---|---|---|
| | 90 - 100 | 33 | 27 | 5 | 0.018 | 0.982 |
| | > 100 | 1 | 1 | 0 | 0.000 | 1.000 |
| Center-forward | | | | | | |
| | 20 - 30 | 465 | 6 | 0 | 0.987 | 0.013 |
| | 30 - 40 | 459 | 16 | 0 | 0.953 | 0.047 |
| | 40 - 50 | 443 | 16 | 0 | 0.918 | 0.082 |
| | 50 - 60 | 427 | 39 | 16 | 0.833 | 0.167 |
| | 60 - 70 | 372 | 49 | 105 | 0.705 | 0.295 |
| | 70 - 80 | 218 | 70 | 71 | 0.435 | 0.565 |
| | 80 - 90 | 77 | 40 | 27 | 0.161 | 0.839 |
| | 90 - 100 | 10 | 9 | 1 | 0.009 | 0.992 |
| Forward (other) | | | | | | |
| | 20 - 30 | 852 | 2 | 0 | 0.998 | 0.002 |
| | 30 - 40 | 850 | 8 | 0 | 0.988 | 0.012 |
| | 40 - 50 | 842 | 22 | 0 | 0.962 | 0.038 |
| | 50 - 60 | 820 | 43 | 16 | 0.912 | 0.089 |
| | 60 - 70 | 761 | 121 | 82 | 0.758 | 0.242 |
| | 70 - 80 | 558 | 198 | 88 | 0.466 | 0.534 |
| | 80 - 90 | 272 | 170 | 43 | 0.150 | 0.850 |
| | 90 - 100 | 59 | 46 | 12 | 0.020 | 0.980 |
| | > 100 | 1 | 1 | 0 | 0.000 | 1.000 |

| Position | Interval | European Cup (1956–1991) N = 2,710 | | | | |
|---|---|---|---|---|---|---|
| | | At risk | Deaths | Censored | Cum. survival prob. | Interval mortality risk |
| Goalkeeper | | | | | | |
| | 20 - 30 | 225 | 1 | 0 | 0.996 | 0.004 |
| | 30 - 40 | 224 | 2 | 0 | 0.987 | 0.013 |
| | 40 - 50 | 222 | 4 | 0 | 0.969 | 0.031 |
| | 50 - 60 | 218 | 15 | 3 | 0.902 | 0.098 |
| | 60 - 70 | 200 | 13 | 43 | 0.836 | 0.164 |
| | 70 - 80 | 144 | 42 | 40 | 0.553 | 0.447 |
| | 80 - 90 | 62 | 28 | 26 | 0.237 | 0.763 |
| | 90 - 100 | 8 | 4 | 4 | 0.079 | 0.921 |
| | > 100 | | | | | |
| Center-back | | | | | | |
| | 20 - 30 | 353 | 3 | 0 | 0.992 | 0.008 |
| | 30 - 40 | 350 | 3 | 0 | 0.983 | 0.017 |
| | 40 - 50 | 347 | 5 | 0 | 0.969 | 0.031 |
| | 50 - 60 | 342 | 14 | 8 | 0.929 | 0.071 |
| | 60 - 70 | 320 | 39 | 71 | 0.801 | 0.199 |
| | 70 - 80 | 210 | 59 | 64 | 0.536 | 0.464 |
| | 80 - 90 | 87 | 47 | 31 | 0.184 | 0.816 |
| | 90 - 100 | 9 | 5 | 4 | 0.053 | 0.948 |
| | > 100 | | | | | |
| Defender (other) | | | | | | |
| | 20 - 30 | 546 | 1 | 0 | 0.998 | 0.002 |
| | 30 - 40 | 545 | 3 | 0 | 0.993 | 0.007 |
| | 40 - 50 | 542 | 6 | 0 | 0.982 | 0.018 |
| | 50 - 60 | 536 | 23 | 18 | 0.939 | 0.061 |
| | 60 - 70 | 495 | 33 | 150 | 0.865 | 0.135 |
| | 70 - 80 | 312 | 72 | 130 | 0.613 | 0.387 |
| | 80 - 90 | 110 | 61 | 43 | 0.191 | 0.810 |
| | 90 - 100 | 6 | 3 | 3 | 0.064 | 0.937 |
| Midfielder | | | | | | |
| | 20 - 30 | 836 | 3 | 0 | 0.996 | 0.004 |
| | 30 - 40 | 833 | 5 | 0 | 0.990 | 0.010 |
| | 40 - 50 | 828 | 11 | 0 | 0.977 | 0.023 |
| | 50 - 60 | 817 | 46 | 42 | 0.921 | 0.079 |
| | 60 - 70 | 729 | 51 | 206 | 0.846 | 0.154 |
| | 70 - 80 | 472 | 90 | 179 | 0.647 | 0.353 |
| | 80 - 90 | 203 | 91 | 83 | 0.282 | 0.718 |
| | 90 - 100 | 29 | 13 | 16 | 0.108 | 0.892 |
| | > 100 | | | | | |
| Center-forward | | | | | | |
| | 20 - 30 | 318 | 2 | 0 | 0.994 | 0.006 |

| | | | | | | |
|---|---|---|---|---|---|---|
| | 30 - 40 | 316 | 4 | 0 | 0.981 | 0.019 |
| | 40 - 50 | 312 | 7 | 0 | 0.959 | 0.041 |
| | 50 - 60 | 305 | 12 | 10 | 0.921 | 0.079 |
| | 60 - 70 | 283 | 33 | 73 | 0.798 | 0.203 |
| | 70 - 80 | 177 | 39 | 59 | 0.587 | 0.413 |
| | 80 - 90 | 79 | 38 | 33 | 0.230 | 0.770 |
| | 90 - 100 | 8 | 6 | 2 | 0.033 | 0.967 |
| Forward (other) | | | | | | |
| | 20 - 30 | 432 | 4 | 0 | 0.991 | 0.009 |
| | 30 - 40 | 428 | 7 | 0 | 0.975 | 0.026 |
| | 40 - 50 | 421 | 5 | 0 | 0.963 | 0.037 |
| | 50 - 60 | 416 | 14 | 11 | 0.930 | 0.070 |
| | 60 - 70 | 391 | 33 | 82 | 0.842 | 0.158 |
| | 70 - 80 | 276 | 65 | 90 | 0.605 | 0.395 |
| | 80 - 90 | 121 | 54 | 53 | 0.259 | 0.741 |
| | 90 - 100 | 14 | 6 | 7 | 0.111 | 0.889 |
| | > 100 | 1 | 1 | 0 | 0.000 | 1.000 |

**Appendix 6**. Cox regressions estimates (with robust standard errors and two-sided tests).

| | World Cup (1930–1990) | | | |
|---|---|---|---|---|
| | Hazard ratio (95% CI) | P value | Hazard ratio (95% CI) | P value |
| Midfielder (reference) | | | | |
| Goalkeeper | 0.73 (0.63–0.84) | <0.001 | 0.73 (0.63–0.84) | <0.001 |
| Center-back | 1.13 (0.97–1.30) | 0.109 | 1.13 (0.98–1.31) | 0.095 |
| Defender (other) | 0.96 (0.83–1.10) | 0.526 | 0.97 (0.84–1.11) | 0.613 |
| Center-forward | 1.28 (1.09–1.50) | 0.003 | 1.27 (1.08–1.50) | 0.005 |
| Forward (other) | 0.95 (0.85–1.06) | 0.394 | 0.95 (0.85–1.06) | 0.350 |
| Northwestern Europe (reference) | | | | |
| South / Central America | | | 1.03 (0.93–1.14) | 0.599 |
| Central / Eastern Europe | | | 1.30 (1.17–1.46) | <0.001 |
| Asia | | | 1.39 (0.97–1.99) | 0.070 |
| Sub-Saharan Africa | | | 2.33 (1.46–3.73) | <0.001 |
| Southern Europe | | | 0.94 (0.81–1.08) | 0.361 |
| Maghreb / Middle East | | | 0.83 (0.63–1.10) | 0.201 |

| | European Cup (1956–1991) | | | |
|---|---|---|---|---|
| | Hazard ratio (95% CI) | P value | Hazard ratio (95% CI) | P value |
| Midfielder (reference) | | | | |
| Goalkeeper | 1.11 (0.88–1.40) | 0.397 | 1.11 (0.88–1.39) | 0.389 |
| Center-back | 1.31 (1.08–1.57) | 0.005 | 1.28 (1.07–1.55) | 0.009 |
| Defender (other) | 1.20 (1.01–1.42) | 0.036 | 1.20 (1.02–1.42) | 0.032 |
| Center-forward | 1.19 (0.97–1.46) | 0.093 | 1.19 (0.97–1.46) | 0.098 |
| Forward (other) | 1.00 (0.83–1.20) | 0.979 | 0.99 (0.82–1.19) | 0.878 |
| Northwestern Europe (reference) | | | | |
| Central / Eastern Europe | | | 1.30 (1.17–1.46) | <0.001 |
| Southern Europe | | | 0.94 (0.81–1.08) | 0.361 |
| non-Europe | | | 0.83 (0.63–1.10) | 0.201 |

*Notes*. Robust standard errors. Two-sided significance tests. Birth cohort stratified. World Cup, full model: 4 223 subjects, 2 333 deaths, time at risk = 301,828, Wald $\chi^2$ (11) = 82.28 (p<0.001). European Cup: 2 710 subjects, 1 126 deaths, time at risk = 194,910, Wald $\chi^2$ (8) = 30.75 (p<0.001). Results reported in Table 2.

**Appendix 7**. Cox regressions estimates with alternative reference categories.

| | World Cup (1930–1990) | | | | | |
|---|---|---|---|---|---|---|
| | Reported (Table 2) | | Goalkeeper reference | | Forward reference | |
| Position | Hazard ratio (95% CI) | P value | Hazard ratio (95% CI) | P value | Hazard ratio (95% CI) | P value |
| Goalkeeper | 0.73 (0.63–0.84) | <0.001 | *ref.* | | 0.77 (0.67–0.89) | <0.001 |
| Center-back | 1.13 (0.98–1.31) | 0.095 | 1.55 (1.31–1.81) | <0.001 | 1.19 (1.03–1.38) | 0.017 |
| Defender (other) | 0.97 (0.84–1.11) | 0.613 | 1.32 (1.12–1.56) | 0.001 | 1.02 (0.89–1.17) | 0.802 |
| Midfielder | *ref.* | | 1.37 (1.19–1.58) | <0.001 | 1.05 (0.94–1.18) | 0.350 |
| Center-forward | 1.27 (1.08–1.50) | 0.005 | 1.74 (1.44–2.10) | <0.001 | 1.34 (1.13–1.58) | 0.001 |
| Forward (other) | 0.95 (0.85–1.06) | 0.350 | 1.30 (1.13–1.50) | <0.001 | *ref.* | |

| | European Cup (1956–1991) | | | | | |
|---|---|---|---|---|---|---|
| | Reported (Table 2) | | Goalkeeper reference | | Forward reference | |
| Position | Hazard ratio (95% CI) | P value | Hazard ratio (95% CI) | P value | Hazard ratio (95% CI) | P value |
| Goalkeeper | 1.11 (0.88–1.39) | 0.389 | *ref.* | | 1.12 (0.87–1.44) | 0.365 |
| Center-back | 1.28 (1.07–1.55) | 0.009 | 1.16 (0.90–1.49) | 0.243 | 1.30 (1.06–1.61) | 0.014 |
| Defender (other) | 1.20 (1.02–1.42) | 0.032 | 1.09 (0.86–1.37) | 0.497 | 1.22 (1.00–1.48) | 0.046 |
| Midfielder | *ref.* | | 0.90 (0.72–1.14) | 0.389 | 1.01 (0.84–1.22) | 0.878 |
| Center-forward | 1.19 (0.97–1.46) | 0.098 | 1.08 (0.83–1.40) | 0.589 | 1.21 (0.96–1.52) | 0.107 |
| Forward (other) | 0.99 (0.82–1.19) | 0.878 | 0.89 (0.69–1.14) | 0.365 | *ref.* | |

*Notes*. Robust standard errors. Two-sided significance tests. Models include the region of origin control variable (not shown). Birth cohort stratified. World Cup, full model: 4 223 subjects, 2 333 deaths, time at risk = 301,828, Wald $\chi^2$ (11) = 82.28 (p<0.001). European Cup: 2 710 subjects, 1 126 deaths, time at risk = 194,910, Wald $\chi^2$ (8) = 30.75 (p<0.001).

The results using alternative reference categories are consistent with adjusted hazard ratios reported in Table 2, as well as the heterogeneity analysis (Figure 3). Relative to goalkeepers, all outfield players in the World Cup cohort display a higher mortality risk, which is not the case in the European Cup cohort. Relative to other forwards, center-backs and center forwards in the World Cup cohort, and center-backs and other defenders in the European cohort, display higher mortality risk.

**Appendix 8**. Position-line-specific survival (World Cup cohort).

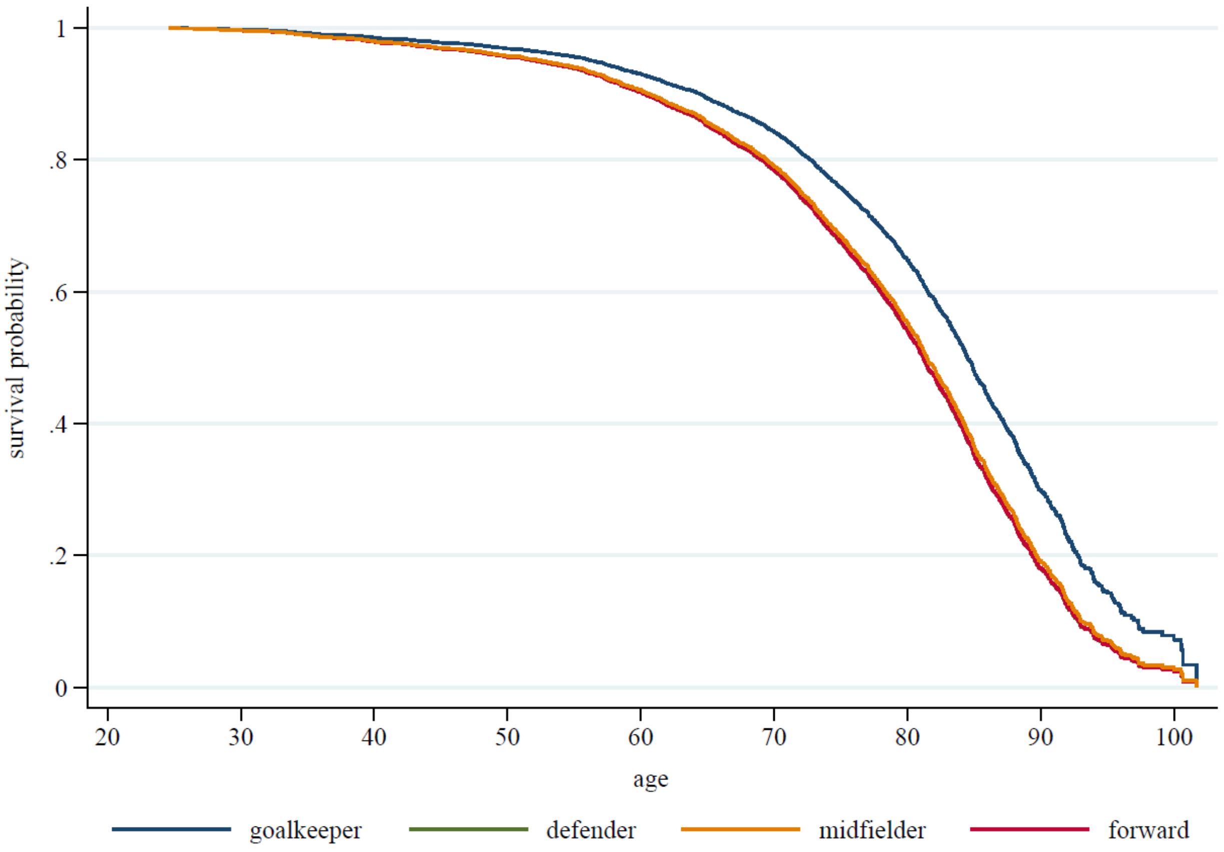

*Notes*. Drawn from a Cox regression model controlling for birth cohort and geographical region, using a crude position *line* variable. Main results refer to detailed positions (Figure 2), reported in Table 2.

**Appendix 9**. Restricted mean survival times (RMST).

| | World Cup (1930–1990) | | | | | | | | | | | | | | |
|---|---|---|---|---|---|---|---|---|---|---|---|---|---|---|---|
| | age 60 | | | age 65 | | | age 70 | | | age 75 | | | age 80 | | |
| Position | RMST | years lost | Δ to MF | RMST | years lost | Δ to MF | RMST | years lost | Δ to MF | RMST | years lost | Δ to MF | RMST | years lost | Δ to MF |
| Goalkeeper | 59.2 | 0.8 | 0.3 | 63.7 | 1.3 | 0.4 | 68.0 | 2.0 | 0.7 | 72.0 | 3.0 | 1.0 | 75.4 | 4.6 | 1.4 |
| Center-back | 58.8 | 1.2 | -0.1 | 63.1 | 1.9 | -0.2 | 67.0 | 3.0 | -0.3 | 70.6 | 4.4 | -0.4 | 73.5 | 6.6 | -0.6 |
| Defender (other) | 58.9 | 1.1 | 0.0 | 63.3 | 1.7 | 0.1 | 67.4 | 2.6 | 0.1 | 71.1 | 3.9 | 0.1 | 74.2 | 5.8 | 0.2 |
| Midfielder | 58.9 | 1.1 | | 63.3 | 1.7 | | 67.3 | 2.7 | | 71.0 | 4.0 | | 74.0 | 6.0 | |
| Center-forward | 58.6 | 1.4 | -0.3 | 62.8 | 2.2 | -0.5 | 66.7 | 3.4 | -0.7 | 70.0 | 5.0 | -1.0 | 72.7 | 7.3 | -1.4 |
| Forward (other) | 58.9 | 1.1 | 0.1 | 63.3 | 1.7 | 0.1 | 67.4 | 2.6 | 0.1 | 71.1 | 3.9 | 0.2 | 74.3 | 5.7 | 0.2 |

| | European Cup (1956–1991) | | | | | | | | | | | | | | |
|---|---|---|---|---|---|---|---|---|---|---|---|---|---|---|---|
| | age 60 | | | age 65 | | | age 70 | | | age 75 | | | age 80 | | |
| Position | RMST | years lost | Δ to MF | RMST | years lost | Δ to MF | RMST | years lost | Δ to MF | RMST | years lost | Δ to MF | RMST | years lost | Δ to MF |
| Goalkeeper | 59.2 | 0.8 | -0.1 | 63.8 | 1.3 | -0.1 | 68.1 | 1.9 | -0.2 | 72.1 | 2.9 | -0.2 | 75.5 | 4.5 | -0.3 |
| Center-back | 59.1 | 0.9 | -0.2 | 63.5 | 1.5 | -0.3 | 67.8 | 2.2 | -0.5 | 71.6 | 3.4 | -0.7 | 74.8 | 5.2 | -1.0 |
| Defender (other) | 59.1 | 0.9 | -0.1 | 63.6 | 1.4 | -0.2 | 67.9 | 2.1 | -0.4 | 71.8 | 3.2 | -0.5 | 75.1 | 4.9 | -0.8 |
| Midfielder | 59.3 | 0.7 | | 63.9 | 1.2 | | 68.3 | 1.8 | | 72.3 | 2.7 | | 75.8 | 4.2 | |
| Center-forward | 59.1 | 0.9 | -0.1 | 63.6 | 1.4 | -0.2 | 67.9 | 2.1 | -0.3 | 71.8 | 3.2 | -0.5 | 75.1 | 4.9 | -0.7 |
| Forward (other) | 59.3 | 0.7 | 0.0 | 63.9 | 1.1 | 0.0 | 68.3 | 1.7 | 0.0 | 72.3 | 2.7 | 0.0 | 75.9 | 4.1 | 0.0 |

**Appendix 10**. Cox regression interaction model estimates.

| | World Cup (1930–1990) | |
| --- | --- | --- |
| | Hazard ratio (95% CI) | P value |
| Midfielder (reference) | | |
| Goalkeeper | 0.78 (0.62–0.96) | 0.021 |
| Center-back | 1.32 (1.06–1.62) | 0.012 |
| Defender (other) | 1.05 (0.85–1.31) | 0.638 |
| Center-forward | 1.58 (1.25–2.00) | <0.001 |
| Forward (other) | 1.08 (0.91–1.28) | 0.368 |
| Northwestern Europe (reference) | | |
| South / Central America | 1.24 (1.01–1.52) | 0.041 |
| Central / Eastern Europe | 1.44 (1.62–1.78) | 0.001 |
| Asia | 1.20 (0.59–2.45) | 0.617 |
| Sub-Saharan Africa | 1.92 (0.80–4.36) | 0.142 |
| Southern Europe | 1.24 (0.98–1.57) | 0.069 |
| Maghreb / Middle East | 1.08 (0.57–2.04) | 0.822 |
| Position (midfielder) × region (Northwestern Europe) | | |
| Goalkeeper × South / Central America | 0.86 (0.59–1.25) | 0.435 |
| Goalkeeper × Central / Eastern Europe | 0.99 (0.67–1.45) | 0.944 |
| Goalkeeper × Asia | 1.63 (0.48–5.55) | 0.432 |
| Goalkeeper × Sub-Saharan Africa | 3.37 (1.03–11.01) | 0.044 |
| Goalkeeper × Southern Europe | 0.63 (0.40–1.00) | 0.048 |
| Goalkeeper × Maghreb / Middle East | 1.14 (0.46–2.84) | 0.784 |
| Center-back × South / Central America | 0.69 (0.47–1.02) | 0.064 |
| Center-back × Central / Eastern Europe | 0.93 (0.66–1.32) | 0.689 |
| Center-back × Asia | 0.67 (0.21–2.09) | 0.492 |
| Center-back × Sub-Saharan Africa | 1.58 (0.28–8.78) | 0.604 |
| Center-back × Southern Europe | 0.73 (0.46–1.17) | 0.195 |
| Center-back × Maghreb / Middle East | 0.59 (0.16–2.17) | 0.429 |
| Defender (other) × South / Central America | 0.87 (0.61–1.23) | 0.425 |
| Defender (other) × Central / Eastern Europe | 1.10 (0.77–1.57) | 0.616 |
| Defender (other) × Asia | 0.77 (0.29–2.07) | 0.611 |
| Defender (other) × Sub-Saharan Africa | 1.67 (0.52–5.39) | 0.389 |
| Defender (other) × Southern Europe | 0.64 (0.40–1.03) | 0.063 |
| Defender (other) × Maghreb / Middle East | 0.53 (0.22–1.24) | 0.144 |
| Center-forward × South / Central America | 0.67 (0.45–1.00) | 0.049 |
| Center-forward × Central / Eastern Europe | 0.75 (0.47–1.21) | 0.244 |
| Center-forward × Asia | 1.46 (0.27–7.92) | 0.660 |
| Center-forward × Sub-Saharan Africa | 0.37 (0.05–2.76) | 0.333 |
| Center-forward × Southern Europe | 0.62 (0.37–1.03) | 0.066 |

| | | |
|---|---|---|
| Center-forward × Maghreb / Middle East | 1.40 (0.53–3.71) | 0.493 |
| Forward (other) × South / Central America | 0.79 (0.60–1.06) | 0.112 |
| Forward (other) × Central / Eastern Europe | 0.79 (0.59–1.06) | 0.121 |
| Forward (other) × Asia | 2.13 (0.87–5.22) | 0.096 |
| Forward (other) × Sub-Saharan Africa | 1.13 (0.20–6.54) | 0.891 |
| Forward (other) × Southern Europe | 0.75 (0.51–1.10) | 0.142 |
| Forward (other) × Maghreb / Middle East | 0.66 (0.30–1.43) | 0.292 |

| | European Cup (1956–1991) | |
|---|---|---|
| | Hazard ratio (95% CI) | P value |
| Midfielder (reference) | | |
| Goalkeeper | 1.28 (0.91–1.78) | 0.153 |
| Center-back | 1.38 (1.06–1.79) | 0.017 |
| Defender (other) | 1.22 (0.96–1.56) | 0.104 |
| Center-forward | 1.29 (0.97–1.72) | 0.081 |
| Forward (other) | 1.09 (0.84–1.41) | 0.533 |
| Northwestern Europe (reference) | | |
| Central / Eastern Europe | 1.43 (1.07–1.89) | 0.014 |
| Non-Europe | 0.84 (0.51–1.37) | 0.484 |
| Southern Europe | 1.09 (0.83–1.43) | 0.533 |
| Position (midfielder) × region (Northwestern Europe) | | |
| Goalkeeper × Southern Europe | 0.63 (0.35–1.12) | 0.112 |
| Goalkeeper × Central / Eastern Europe | 0.96 (0.55–1.69) | 0.897 |
| Goalkeeper × non-Europe | 0.67 (0.24–1.85) | 0.435 |
| Center-back × Southern Europe | 0.70 (0.42–1.16) | 0.162 |
| Center-back × Central / Eastern Europe | 0.98 (0.63–1.52) | 0.916 |
| Center-back × non-Europe | 1.13 (0.50–2.57) | 0.763 |
| Defender (other) × Southern Europe | 0.96 (0.64–1.45) | 0.864 |
| Defender (other) × Central / Eastern Europe | 0.98 (0.65–1.49) | 0.941 |
| Defender (other) × non-Europe | 0.79 (0.32–1.95) | 0.611 |
| Center-forward × Southern Europe | 1.10 (0.65–1.85) | 0.722 |
| Center-forward × Central / Eastern Europe | 0.77 (0.44–1.32) | 0.340 |
| Center-forward × non-Europe | 0.67 (0.30–1.52) | 0.337 |
| Forward (other) × Southern Europe | 0.82 (0.50–1.36) | 0.441 |
| Forward (other) × Central / Eastern Europe | 0.70 (0.44–1.12) | 0.139 |
| Forward (other) × non-Europe | 1.53 (0.74–3.16) | 0.248 |

*Notes*. Robust standard errors. Two-sided significance tests. Birth cohort stratified. World Cup: 4 223 subjects, 2 333 deaths, time at risk = 301,828, Wald $\chi^2$ (41) = 141.23 (p<0.001). European Cup: 2 710 subjects, 1 126 deaths, time at risk = 194,910, Wald $\chi^2$ (23) = 47.67 (p=0.002). Results reported in Figure 3.

**Appendix 11**. Marginal hazard rations from Cox regression interaction model estimates.

| World Cup (1930–1990) | |
|---|---|
| | Adjusted Hazard ratio (95% CI) |
| Goalkeeper × South / Central America | 0.83 (0.59–1.06) |
| Goalkeeper × Northwestern Europe | 0.78 (0.61–0.94) |
| Goalkeeper × Central / Eastern Europe | 1.10 (0.78–1.41) |
| Goalkeeper × Asia | 1.52 (0.02–3.01) |
| Goalkeeper × Sub-Saharan Africa | 5.03 (1.04–9.03) |
| Goalkeeper × Southern Europe | 0.61 (0.39–0.83) |
| Goalkeeper × Maghreb / Middle East | 0.95 (0.34–1.56) |
| Center-back × South / Central America | 1.13 (0.79–1.47) |
| Center-back × Northwestern Europe | 1.32 (1.03–1.60) |
| Center-back × Central / Eastern Europe | 1.76 (1.33–2.19) |
| Center-back × Asia | 1.06 (0.13–1.99) |
| Center-back × Sub-Saharan Africa | 3.99 (−1.91–9.90) |
| Center-back × Southern Europe | 1.20 (0.74–1.66) |
| Center-back × Maghreb / Middle East | 0.84 (−0.10–1.78) |
| Defender (other) × South / Central America | 1.13 (0.85–1.41) |
| Defender (other) × Northwestern Europe | 1.05 (0.83–1.28) |
| Defender (other) × Central / Eastern Europe | 1.66 (1.23–2.08) |
| Defender (other) × Asia | 0.98 (0.33–1.63) |
| Defender (other) × Sub-Saharan Africa | 3.39 (0.77–6.02) |
| Defender (other) × Southern Europe | 0.84 (0.52–1.16) |
| Defender (other) × Maghreb / Middle East | 0.60 (0.27–0.93) |
| Midfielder × South / Central America | 1.24 (0.98–1.49) |
| Midfielder × Northwestern Europe | *ref.* |
| Midfielder × Central / Eastern Europe | 1.44 (1.13–1.74) |
| Midfielder × Asia | 1.20 (0.34–2.06) |
| Midfielder × Sub-Saharan Africa | 1.92 (0.24–3.61) |
| Midfielder × Southern Europe | 1.24 (0.95–1.54) |
| Midfielder × Maghreb / Middle East | 1.08 (0.39–1.76) |
| Center-forward × South / Central America | 1.30 (0.91–1.70) |
| Center-forward × Northwestern Europe | 1.58 (1.20–1.95) |
| Center-forward × Central / Eastern Europe | 1.70 (1.03–2.38) |
| Center-forward × Asia | 2.77 (−1.45–6.98) |
| Center-forward × Sub-Saharan Africa | 1.13 (−0.90–3.16) |
| Center-forward × Southern Europe | 1.22 (0.71–1.73) |
| Center-forward × Maghreb / Middle East | 2.38 (0.65–4.12) |
| Forward (other) × South / Central America | 1.06 (0.85–1.27) |
| Forward (other) × Northwestern Europe | 1.08 (0.90–1.26) |
| Forward (other) × Central / Eastern Europe | 1.23 (0.98–1.48) |
| Forward (other) × Asia | 2.77 (1.29–4.25) |
| Forward (other) × Sub-Saharan Africa | 2.35 (−1.24–5.95) |
| Forward (other) × Southern Europe | 1.01 (0.70–1.31) |
| Forward (other) × Maghreb / Middle East | 0.77 (0.43–1.10) |

| European Cup (1956–1991) | |
| --- | --- |
| | Adjusted Hazard ratio (95% CI) |
| Goalkeeper × Northwestern Europe | 1.28 (0.85–1.70) |
| Goalkeeper × Southern Europe | 0.87 (0.48–1.26) |
| Goalkeeper × Central / Eastern Europe | 1.75 (1.02–2.49) |
| Goalkeeper × non-Europe | 0.71 (0.09–1.34) |
| Center-back × Northwestern Europe | 1.38 (1.02–1.74) |
| Center-back × Southern Europe | 1.05 (0.62–1.48) |
| Center-back × Central / Eastern Europe | 1.92 (1.31–2.53) |
| Center-back × Central / Eastern Europe | 1.31 (0.47–2.16) |
| Defender (other) × Northwestern Europe | 1.22 (0.93–1.52) |
| Defender (other) × Southern Europe | 1.29 (0.90–1.67) |
| Defender (other) × Central / Eastern Europe | 1.72 (1.21–2.23) |
| Defender (other) × non-Europe | 0.81 (0.20–1.43) |
| Midfielder × Northwestern Europe | *ref.*. |
| Midfielder × Southern Europe | 1.09 (0.79–1.39) |
| Midfielder × Central / Eastern Europe | 1.43 (1.02–1.83) |
| Midfielder × non-Europe | 0.84 (0.43–1.25) |
| Center-forward × Northwestern Europe | 1.29 (0.92–1.66) |
| Center-forward × Southern Europe | 1.55 (0.91–2.18) |
| Center-forward × Central / Eastern Europe | 1.41 (0.79–2.03) |
| Center-forward × non-Europe | 0.73 (0.27–1.18) |
| Forward (other) × Northwestern Europe | 1.09 (0.80–1.37) |
| Forward (other) × Southern Europe | 0.97 (0.58–1.37) |
| Forward (other) × Central / Eastern Europe | 1.09 (0.71–1.47) |
| Forward (other) × non-Europe | 1.40 (0.67–2.13) |

*Notes*. Estimates drawn from models presented in Figure 3 (appendix 10 pp 24–25), using equation 4. Hazard ratios were estimated using post-estimation marginal predictions from Cox proportional hazards models with position–region interactions. Point estimates correspond to exponentiated linear predictors. Confidence intervals were obtained using the delta method applied to the transformed estimand. For some position-region combinations with small sample sizes (e.g., Asia, Sub-Saharan Africa and Maghreb / Middle East), the resulting confidence intervals may extend below zero due to large estimation uncertainty. Such intervals reflect imprecision rather than negative hazards.
